# Longitudinal Analysis of the Burden of Post-Acute Chikungunya-Associated Arthralgia in Children and Adults: A Prospective Cohort Study in Managua, Nicaragua (2014-2019)

**DOI:** 10.1101/2023.05.09.23289726

**Authors:** Colin M. Warnes, Fausto Andres Bustos Carrillo, Jose Victor Zambrana, Brenda Lopez Mercado, Sonia Arguello, Oscarlette Ampié, Damaris Collado, Nery Sanchez, Sergio Ojeda, Guillermina Kuan, Aubree Gordon, Angel Balmaseda, Eva Harris

## Abstract

Chikungunya can result in debilitating arthralgia, often presenting as acute, self-limited pain, but occasionally manifesting chronically. Little is known about differences in chikungunya-associated arthralgia comparing children to adults over time. To characterize long-term chikungunya-associated arthralgia, we recruited 770 patients (105 0-4 year olds [y/o], 200 5-9 y/o, 307 10-15 y/o, and 158 16+ y/o) with symptomatic chikungunya virus infections in Managua, Nicaragua, during two chikungunya epidemics (2014-2015). Participants were assessed at ∼15 days and 1, 3, 6, 12, and 18 months post-fever onset. Following clinical guidelines, we defined participants by their last reported instance of arthralgia as acute (≤10 days post-fever onset), interim (>10 and <90 days), or chronic (≥90 days) cases. We observed a high prevalence of arthralgia (80-95%) across all ages over the study period. Overall, the odds of acute arthralgia increased in an age-dependent manner, with the lowest odds of arthralgia in the 0-4 y/o group (odds ratio [OR]: 0.27, 95% confidence interval [CI]: 0.14-0.51) and the highest odds of arthralgia in the 16+ y/o participants (OR: 4.91, 95% CI: 1.42-30.95) relative to 10-15 y/o participants. Females had a higher odds of acute arthralgia than males (OR: 1.63, 95% CI: 1.01-2.65) across all ages. We found that 23-36% of pediatric and 53% of adult participants reported an instance of post-acute arthralgia. Children exhibited the highest prevalence of post-acute polyarthralgia in their legs, followed by the hands and torso – a pattern not seen among adult participants. Further, we observed pediatric chikungunya presenting in two distinct phases: the acute phase and the associated interim and chronic phases. Differences in the presentation of arthralgia were observed across age, sex, and disease phase in this longitudinal chikungunya cohort. Our results elucidate the long-term burden of chikungunya-associated arthralgia among pediatric and adult populations.

**Author Summary:** Upon its emergence in the Americas in 2013, chikungunya virus spread rapidly, leading to >2 million suspected autochthonous cases between 2014-2015. Much of what we know about chikungunya is derived from adult populations, leading to gaps in guidelines to treat pediatric chikungunya. To address these gaps, we assembled a large cohort of both pediatric (n=612) and adult (n=158) laboratory-confirmed (n=682) or clinically/epidemiologically probable (n=88) chikungunya cases from two distinct epidemics in 2014 and 2015 in Managua, Nicaragua, followed these patients over a two-year timeline, and analyzed chikungunya-associated arthralgia using rigorous statistical approaches. Our analysis demonstrates that the pediatric (0–15 years old [y/o]) population faces a previously unappreciated high burden of post-acute chikungunya-associated arthralgia. Further, we observe post-acute arthralgia presents differently between pediatric and adult cases (16+ y/o). The difference between the two groups was evident when comparing distribution of polyarthralgia across the body parts and when analyzing the persistence of arthralgia in the post-acute phase (> 10 days post-fever onset). Using detailed longitudinal data, our findings provide insight into long-term chikungunya arthralgia across age, sex, body parts, and the different stages of chikungunya. We believe these findings will inform clinical guidelines regarding chikungunya-associated arthralgia across all ages.

## Introduction

Chikungunya virus (CHIKV) is a mosquito-borne alphavirus that typically presents with high fever, rash, headache, myalgia, and debilitating polyarthralgia (1). Chikungunya is often described as having three phases: acute, post-acute/subacute/interim, and chronic (2). The acute symptoms last 7-10 days post-symptom onset, per the US Centers for Disease Control and Prevention (CDC) (3). Acute chikungunya can present nonspecifically, complicating differential diagnosis (4). Though most clinical manifestations subside after the acute phase, chronic arthralgia has been reported up to 6 years post-infection (5). Chronic chikungunya is often defined as chikungunya-associated sequelae >3 months post-symptom onset (2,6–9). The period between acute and chronic phases has been described as the subacute or post-acute period, which we call the *interim phase* (2,6,8,10–13). Being over 45 years old (y/o) and female are considered risk factors for developing chikungunya-associated symptoms and sequelae (2,6,10). Despite recent studies on the short-and long-term presentation of chikungunya (2,3,6,9,14–17), the literature is limited regarding the pediatric burden of chikungunya-associated arthralgia over time (10,11), a particular problem given that children are a vulnerable population that has been known to experience high morbidity in prior outbreaks (18,19).

In Managua, Nicaragua, CHIKV was first detected in July 2014, with autochthonous transmission observed in September 2014 (20). CHIKV caused two epidemics, a moderately sized epidemic in 2014-2015 and a much larger epidemic in 2015-2016, both caused by the Asian lineage (19,21,22). We assessed the presence of arthralgia for >18 months post-fever onset among a cohort of chikungunya cases that were recruited from the Pediatric Dengue Cohort Study (PDCS), a longitudinal cohort of ∼4000 children in Managua, as well as adults who received medical care from the same health facility, the Health Center Sócrates Flores Vivas (HCSFV). We quantified differences between adult and pediatric cases and characterized the presentation of arthralgia over time to fill existing gaps in our knowledge of long-term pediatric arthralgia (10,14,23–25). Using our cohort, we demonstrate the age dependence of arthralgia occurrence, define the pediatric burden of arthralgia, and describe the characteristics of the acute, interim, and chronic phases of disease.

## Methods

### Ethics statement

This study was approved by the Institutional Review Boards of the University of California, Berkeley, and the Nicaraguan Ministry of Health. Participants aged 18 and older and parents or legal representatives of participating children (0-17 y/o) provided written informed consent. Children 6-17 y/o provided verbal assent.

### Study recruitment

All participants were recruited from the HCSFV during the 2014 and 2015 chikungunya epidemics in Managua. During enrollment, study personnel administered informed consent and recruited participants based on laboratory-confirmed or suspected clinical presentation with chikungunya (see Supplemental Methods). Individuals aged 6 months and older were considered for the study. Inclusion criteria for the study were as follows: 1) laboratory-confirmed or clinically probable CHIKV infection, 2) having been attended to at the HCSFV, 3) residing in the HCSFV catchment area during the study period, 4) having provided written informed consent themselves or through a legal representative if <18 y/o, and 5) having provided verbal assent if between 6 and 17 y/o (3,4). Participants in a parallel Zika cohort followed similar inclusion criteria but with clinical and laboratory diagnostic methods for Zika (see Supplemental Methods).

### Study design

Participants were evaluated upon enrollment and followed up at approximately 15 days and 1, 3, 6, 12, and 18 months post-fever onset. At each study visit, study physicians administered a questionnaire to the participant or legal guardian and conducted a physical examination that queried for arthralgia (26). Arthralgia was defined as verbal or physical signs of joint pain, discomfort, or inflammation. The occurrence of arthralgia was assessed across the neck, shoulders, back, hips, elbows, wrists, hands, knees, ankles, and feet. The study period, covering both chikungunya epidemics and the follow-up visits, extended from September 2014 to January 2018. Pediatric participants 2-14 y/o were co-enrolled in the PDCS (27). Adult and infant (6 months to 2 y/o) participants were recruited from the HCSFV and enrolled based on fulfillment of the inclusion criteria listed above. All participants received care at the HCSFV throughout the study.

### Phases of chikungunya

Participants were classified as experiencing acute, interim, or chronic arthralgia cases of chikungunya. Acute cases were defined as experiencing illness strictly within the first 10 days post-fever onset based on CDC and World Health Organization (WHO) guidelines (3,9). Chronic cases of chikungunya were defined as experiencing chikungunya-associated signs or symptoms ≥90 days post-fever onset based on WHO and French guidelines (2,9). The interim cases covered the period from 11-89 days post-fever onset (2,6). Collectively, the interim and chronic cases constitute the *post-acute* phase of chikungunya, defined as any report of arthralgia >10 days post-fever onset. See Supplemental Methods for additional details.

### Data and statistical analysis

Participants were stratified into four age ranges: 0-4, 5-9, 10-15, and 16+ y/o (Table 1) based on established protocols of the PDCS and the Nicaraguan Ministry of Health. We used logistic regression to estimate odds ratios (ORs) for binary outcomes. Generalized additive models (GAM) were used to visualize the non-linear relationship between different variables (28). Pearson’s Chi-squared test was used to compare the proportion of participants with arthralgia across different criteria (29). Survival data were analyzed using proportional hazards models, Kaplan-Meier curves, log rank tests, and log rank tests for trend (30). Agglomerative hierarchical clustering was used to visualize the co-occurrence of participants’ clinical manifestations (31). See Supplemental Methods for additional details. Data were analyzed using R (v4.1.1).

**Table 1.**
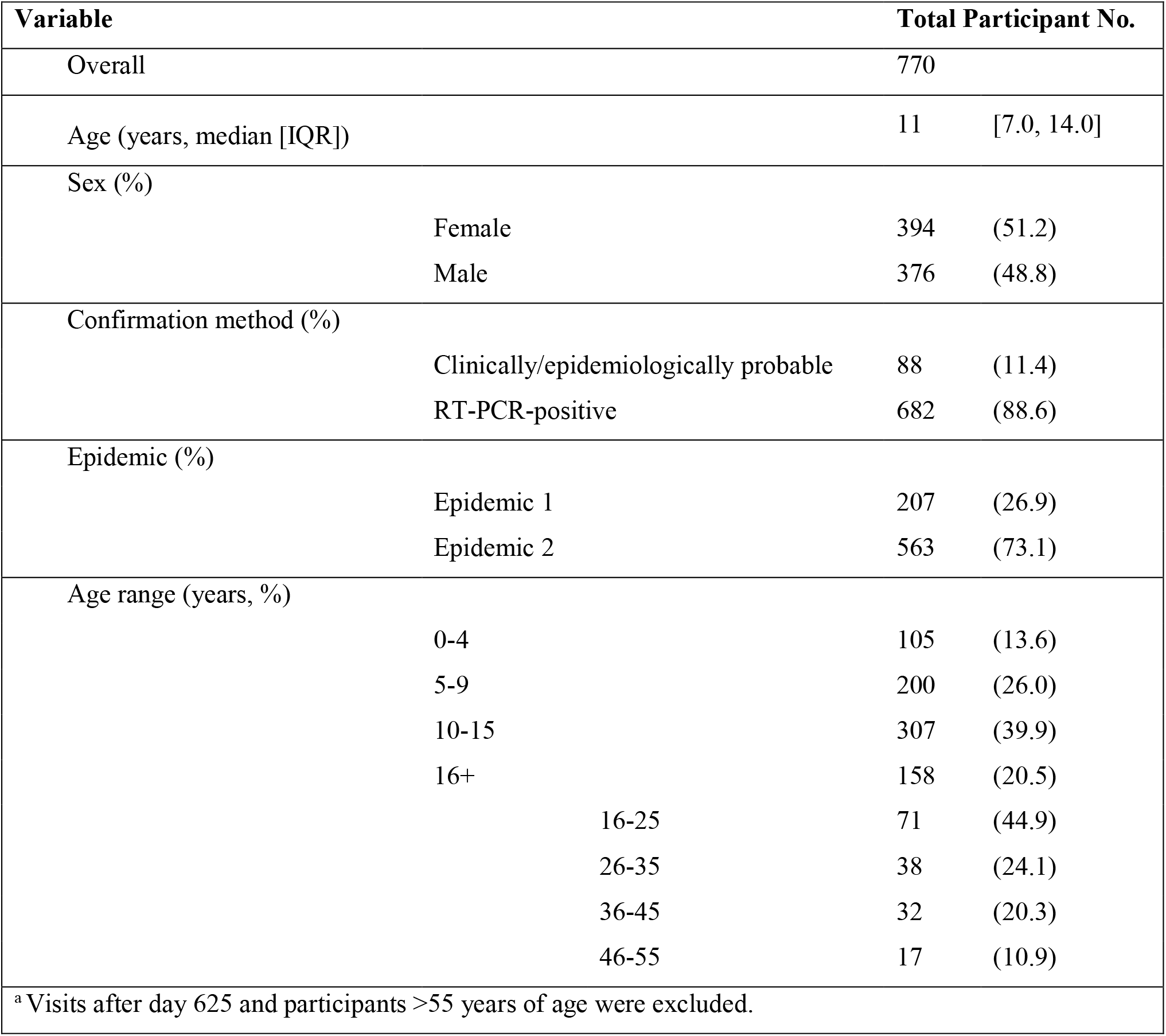
Characteristics of cohort participants.

## Results

### Participant characteristics

Participants consisted of 770 chikungunya cases; 682 (88.6%) were positive by real-time RT-PCR and 88 (11.4%) were clinically/epidemiologically probable cases. Study participants’ median age was 11 y/o (interquartile range: 7-14). We enrolled 394 (51.2%) females and 376 (48.8%) males. There were 612 pediatric (0-15 y/o) and 158 adults (16+ y/o) cases (Table 1). During the chikungunya epidemics, there were very few cases of dengue or Zika – diseases that can present with similar symptoms as chikungunya – detected in the PDCS, limiting the number of incorrectly classified chikungunya cases based on clinical/epidemiological criteria (21).

### Age-based differences in chikungunya-associated arthralgia

We first examined the occurrence of arthralgia across age. Overall, ∼70-95% of pediatric and ∼95-100% of adult participants reported arthralgia during the study period (Fig. S2). We observed that the prevalence of arthralgia increased in an age-dependent manner but decreased over time since symptom onset (Fig. 1). Kaplan-Meier (KM) plots demonstrated an age-dependent increase in percentage of participants with reported post-acute arthralgia across the ∼1.5 year study period (Fig. 2A). The pediatric groups reported ∼ 30% of their total instances of arthralgia occurring beyond the acute phase, showing the substantial burden of chikungunya-associated arthralgia among children (Table S1). Crucially, a large percent of pediatric participants (0-4 y/o: 18.6%; 5-9 y/o: 23.2%; 10-14 y/o: 32.5%) reported 2+ visits with symptoms of arthralgia (Table S2). In contrast, the adult group reported 43.4% of their total instances of arthralgia in the post-acute phase, a significantly higher proportion compared to all other age groups (p-value < 0.05) (Table S1). Adults reported the highest percentage (55.1%) of individuals with 2+ visits with symptoms of arthralgia (Table S2). Age-dependent trends in reported arthralgia occurrence were supported by KM estimates (Fig. 2A), ORs (Table 2), and hazard ratios (HRs) (Table S3).

**Table 2.** Risk factors for acute chikungunya-associated arthralgia.

| Category | Reported Arthralgia |  |  |  | Odds Ratio (95% CI) |  |
| --- | --- | --- | --- | --- | --- | --- |
|  | No | (%) | Yes | ( %) |  |  |
| Age ranges (years) |  |  |  |  |  |  |
| 0-4 | 23 | (22.55) | 79 | (77.5) | 0.27 | (0.14, 0.51) |
| 5-9 | 30 | (15.2) | 168 | (84.8) | 0.44 | (0.24, 0.78) |
| 10-15 | 22 | (7.3) | 280 | (92.7) | - |  |
| 16+ | 2 | (1.6) | 125 | (98.4) | 4.91 | (1.42, 30.95) |
| Sex |  |  |  |  |  |  |
| Female | 31 | (8.3) | 341 | (91.7) | 1.63 | (1.01, 2.65) |
| Male | 46 | (12.9) | 311 | (87.1) | - |  |
| a Some participants were omitted due to missing acute stage data, n = 41 |  |  |  |  |  |  |

**Figure 1.**
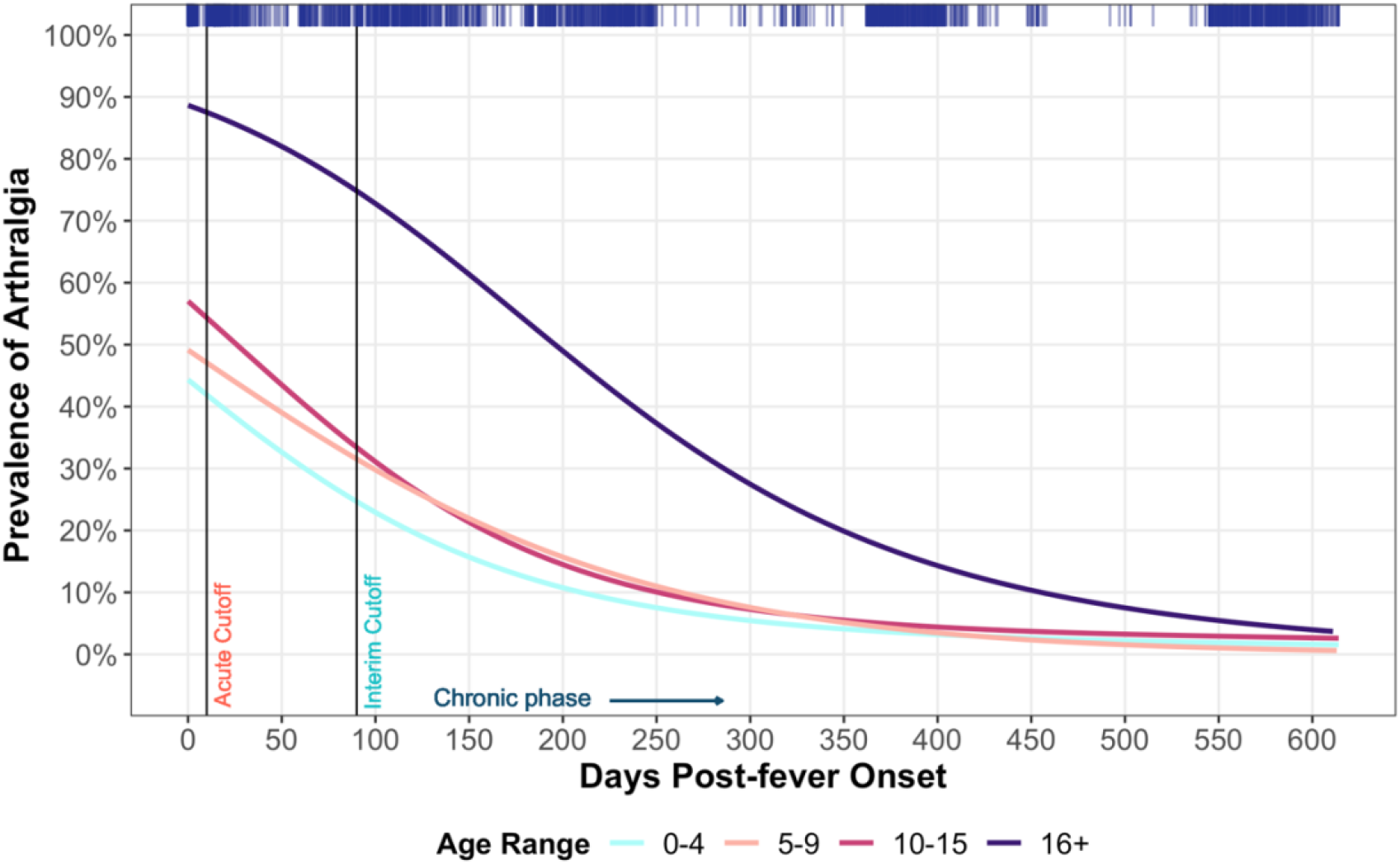
Prevalence of arthralgia over time, stratified by age. The prevalence of arthralgia measured across days since fever onset and stratified by age range is depicted using a GAM model. Top bar indicates the density of patient responses by day since fever onset. Participants were considered as having either acute (≤10 days), interim (>10 and <90 days), or chronic (≥90 days) disease

**Figure 2.**
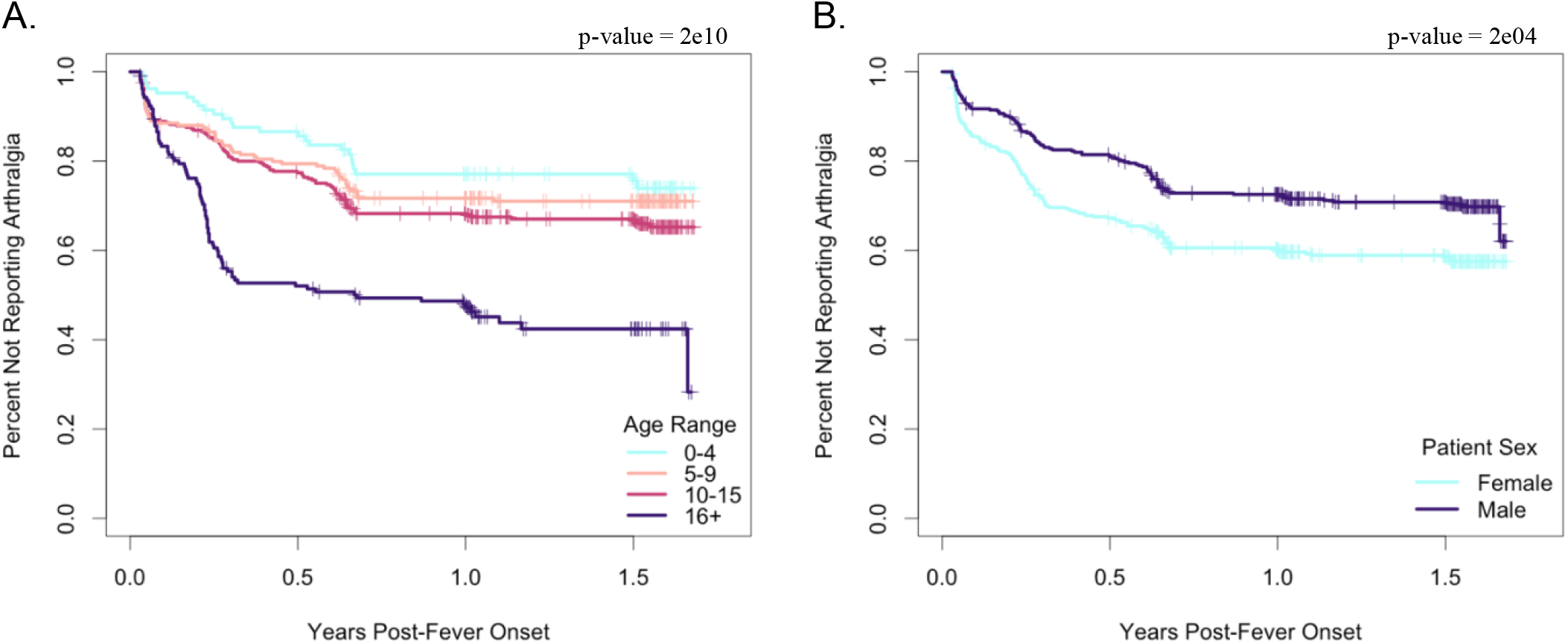
Kaplan-Meier plot showing percent of participants reporting arthralgia over time. A Kaplan-Meier graph plotting percent of participants not reporting arthralgia (y-axis) against years since study entry (x-axis). Ticks correspond to censoring events. Panels show the distribution of participants beginning 10 days post-fever onset and ending at the last reported data point, stratified by age range **(A)** and sex **(B)**.

### Clinical presentation of post-acute chikungunya-associated polyarthralgia

Clustering analyses demonstrated that polyarthralgia co-occurred in three general areas, constituting distinct clusters of localized arthralgia: the legs (knees, ankles, and feet), hands (wrists and hands), and torso/elbows (neck, shoulders, back, hips, and elbows) (Fig. 3A). The most fundamental difference in polyarthralgia co-occurrence was the division of the hand and leg areas, which clustered together, from the torso/elbows, which formed its own cluster.

**Figure 3.**
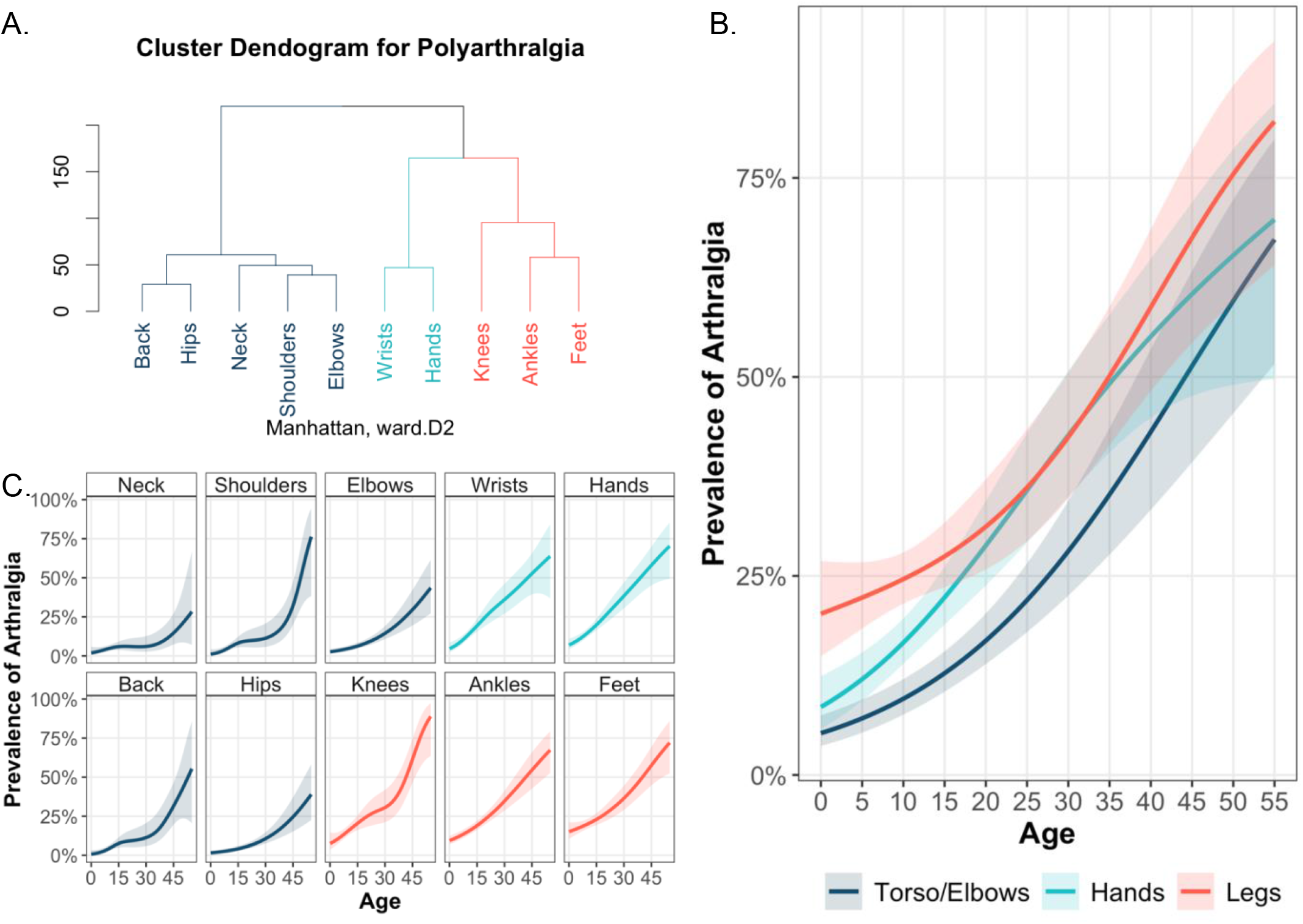
Reported polyarthralgia beyond the acute phase of disease by body part and age. **(A)** Cluster dendrogram depicting the relationship between occurrence of polyarthralgia across the different body parts; cophenetic distance correlation coefficient = 0.95. Age trends of the prevalence of arthralgia among clustered body groups **(B)** and individual body parts **(C)**. Depicted using a GAM model.

The prevalence of arthralgia increased across age for each of the three clusters identified through hierarchical clustering (Fig. 3B). The prevalence of polyarthralgia was highest in the legs (28.5%), followed by the hands (21.1%) and the torso/elbows (13.5%) (Table S4), where the proportion of polyarthralgia between distinct body parts was significantly different in all age groups. Specifically, among pediatric participants (≤15 y/o), the difference between localized polyarthralgia in the hands and legs was significantly different (p-value < 0.001), while this difference was not observed in adults (>15 y/o, p-value = 0.43), indicating distinct presentation of polyarthralgia between the two groups. For each cluster, the prevalence of polyarthralgia increased substantially and nearly linearly over age, particularly after age 20 (Fig 3B). Such trends represent averages across the underlying body parts, which we also characterized: in general, the age-specific prevalence of arthralgia for the individual body parts resembled the trends of the corresponding cluster (Fig. 3C), supporting the clustering analysis and extending its results across age.

### Sex-based differences in chikungunya-associated arthralgia

Over 85% of males and females reported arthralgia over the study period. Over 18 months of follow-up, females experienced significantly higher odds of arthralgia (OR: 1.63 [95% CI: 1.01-2.65]) than males (Table 2). Beyond the acute phase of disease, a significantly higher proportion of females experienced arthralgia compared to males (p < 0.001; log rank test) (Fig. 2B). During this same, the hazard for females experiencing arthralgia was also higher compared to males (HR: 2.27 [95% CI: 1.49-3.46]) (Table S2), both among children (HR: 1.97 [95% CI: 1.18-3.28]) and adults (OR: 2.20 [95% CI: 1.11-4.35]). Sex-based differences in arthralgia occurrence were most apparent within the first six months of follow-up, and this difference remained relatively unchanged until study cessation (Fig. 2B). Altogether, these data suggest distinct, sex-based experiences of arthralgia that are especially pronounced beyond the acute phase.

### Differences across the acute, interim, and chronic phases

We then evaluated the proportion of acute, interim, and chronic arthralgia cases, defined by their last instance of reported arthralgia, across continuous age. We observed that as age increased, the proportion of acute cases decreased while the proportion of interim or chronic cases increased (Fig. 4A). Among younger participants (<15 y/o), ∼20% were considered chronic and ∼10% interim cases, with ∼55% considered acute cases and a subset reporting no arthralgia (∼15%) (Table S5). A high proportion of the older participants were considered interim (26.0%) and chronic cases (29.1%), while fewer were defined as acute cases (43.3%) or did not report arthralgia (1.6%) (Table S5). At ∼18 years of age, the proportion that experienced interim or chronic arthralgia increased up to 50%, with the percentage increasing dramatically as age increased (Fig. 4A). Indeed, by age 30, the proportion of chronic and interim cases was approximately 90%. When comparing only pediatric (<15 y/o) and adult (>15 y/o) participants, we observed that adults had significantly higher interim cases (p-value < 0.001) but not chronic cases (p-value = 0.06). This difference is driven by the significantly lower proportion of post-acute pediatric cases being defined as interim cases (29.4%) rather than chronic cases (70.6%, p-value < 0.001), a difference not observed between interim (47.1%) and chronic (52.9%) cases among adults (p-value = 0.674). Thus, differences in the presentation of arthralgia between the pediatric and adult groups were primarily driven by the occurrence of arthralgia during the interim period.

**Figure 4.**
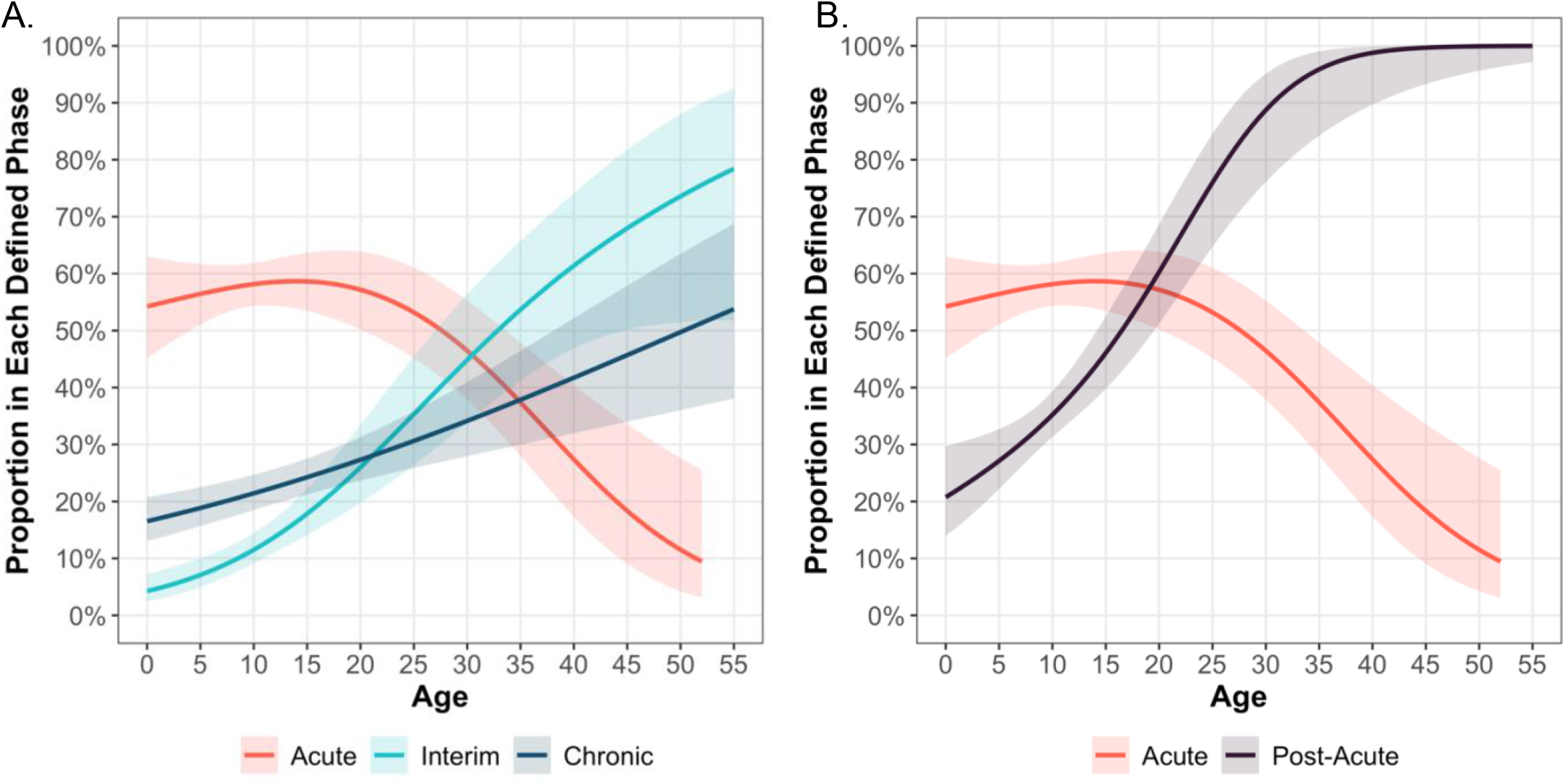
Age trends for the proportion of arthralgia cases. Participants were considered as acute (≤10 days), interim (>10 days and <90 days), or chronic (≥90 days) phase arthralgia cases **(A)** or considered as either acute (≤10 days) or post-acute (>10 days) phase arthralgia cases **(B)**, based on their last instance of arthralgia. The y-axis reflects, out of all participants with reported arthralgia, what proportion had their last instance of arthralgia in each given phase. Depicted using a GAM model.

### Association between the interim and chronic phases

The age-specific trends of interim and chronic arthralgia cases were broadly similar to each other in the pediatric group, but distinct from the trends observed in the adults, suggesting there were differences in the presentation of interim and chronic arthralgia between the two groups (Fig. 4A). To test this hypothesis, we used logistic regression to quantify the association between having either acute or interim arthralgia and later developing chronic arthralgia. Among the pediatric (0-15 y/o) participants, having interim arthralgia was significantly associated with exhibiting chronic arthralgia, and there was no evidence that experiencing acute arthralgia was associated with developing chronic arthralgia (Table 3). Similar results were obtained after adjusting for sex and continuous age. However, among the adult group, the OR for interim arthralgia associated with developing chronic arthralgia was not statistically significant in either the unadjusted or adjusted models. As the regression results strengthened our finding that the interim and chronic phases were similar in the pediatric group, but not adults, we analyzed the proportion of acute and post-acute (combining the interim and chronic cases) arthralgia cases over continuous age (Fig. 4B). The proportion of post-acute cases increased linearly with continuous age, ranging from 25% to 45% among the youngest ages (0-15 y/o) and overtaking the proportion of acute cases around age 20 y/o before reaching 100% by the age of 35 y/o (Fig. 4B). Further, we observed a significant different between the proportion of pediatric and adult patients with post-acute arthralgia (p-value < 0.001). Our analysis suggests that a child experiencing interim arthralgia is likely also to experience chronic phase; however, having interim arthralgia as an adult is not predictive of chronic pain.

**Table 3.**
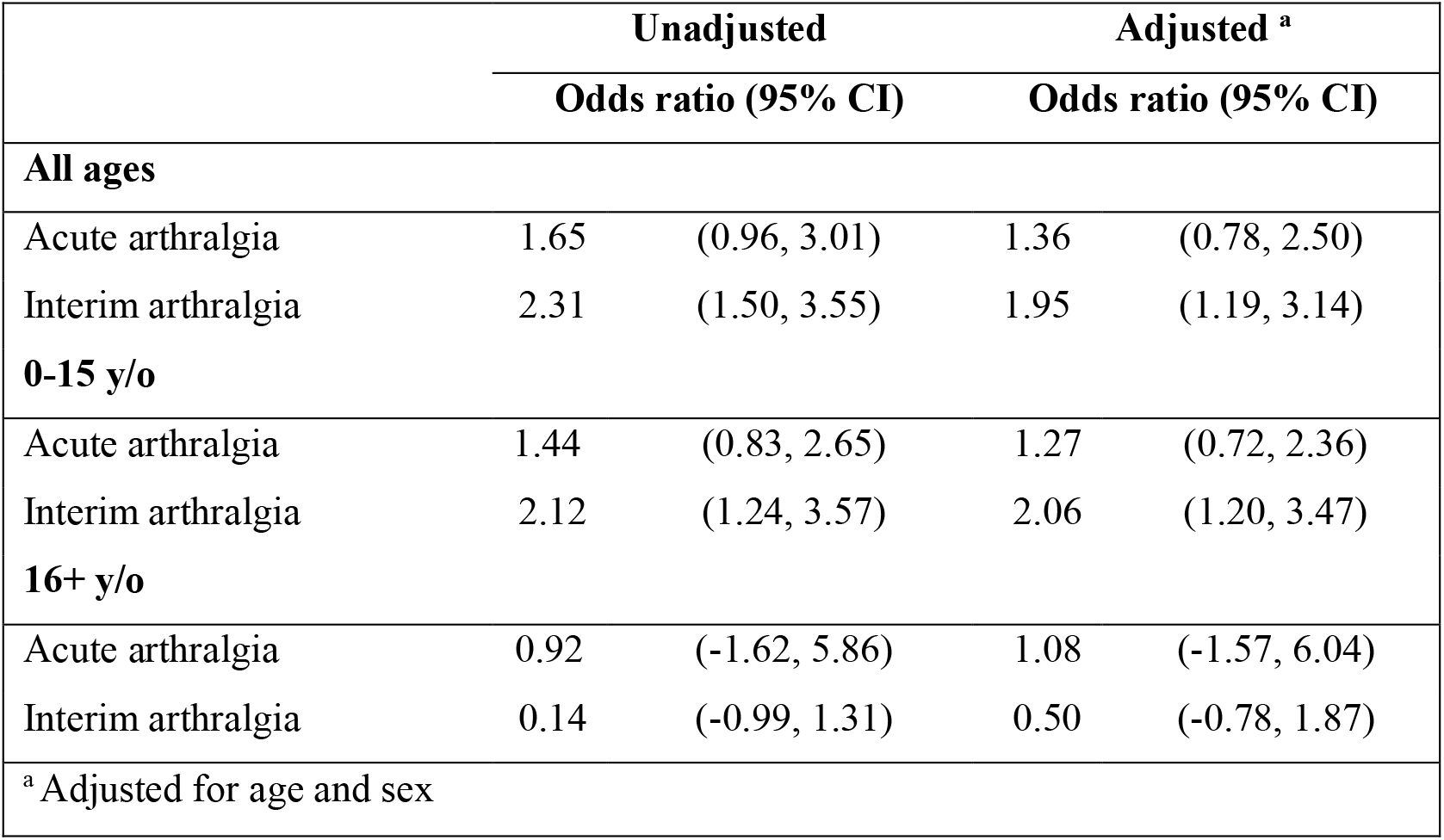
Odds of presenting chronic chikungunya-associated arthralgia.

|  | Unadjusted |  | Adjusted <sup>a</sup> |  |
| --- | --- | --- | --- | --- |
|  | Odds ratio (95% CI) |  | Odds ratio (95% CI) |  |
| All ages |  |  |  |  |
| Acute arthralgia | 1.65 | (0.96, 3.01) | 1.36 | (0.78, 2.50) |
| Interim arthralgia | 2.31 | (1.50, 3.55) | 1.95 | (1.19, 3.14) |
| 0-15 y/o |  |  |  |  |
| Acute arthralgia | 1.44 | (0.83, 2.65) | 1.27 | (0.72, 2.36) |
| Interim arthralgia | 2.12 | (1.24, 3.57) | 2.06 | (1.20, 3.47) |
| 16+ y/o |  |  |  |  |
| Acute arthralgia | 0.92 | (-1.62, 5.86) | 1.08 | (-1.57, 6.04) |
| Interim arthralgia | 0.14 | (-0.99, 1.31) | 0.50 | (-0.78, 1.87) |
| <sup>a</sup> Adjusted for age and sex |  |  |  |  |

### Sensitivity analysis

As a final analysis, we examined whether the confirmation method or baseline levels of pediatric arthralgia impacted our major results. RT-PCR-confirmed and clinically/epidemiologically confirmed participants showed no differences in reported arthralgia occurrence over time (Fig. S3A). However, the chikungunya cohort reported a significantly higher percentage of arthralgia occurrence beyond the acute phase of disease compared to the parallel Zika pediatric cohort, consistent with known clinical differences between chikungunya and Zika. This demonstrates that the levels of arthralgia observed in the cohort were distinct from baseline pediatric arthralgia (Fig. S3B).

## Discussion

We describe risk factors and characteristics of long-term arthralgia among 770 chikungunya cases in a prospective cohort in Managua, Nicaragua. We find that pediatric chikungunya cases are most vulnerable to chikungunya-associated arthralgia during the acute phase, but they do exhibit a meaningfully high prevalence of arthralgia occurring beyond the acute phase of disease. Further, we demonstrate an age-dependent increase in the prevalence of arthralgia from infancy throughout adulthood. While the association between age and arthralgia has been described in adults (14), we present the first comprehensive analysis of pediatric chikungunya-associated arthralgia (11). The highest prevalence of post-acute pediatric polyarthralgia was reported in the legs, followed by the hands and torso/elbows; no significant differences among body parts was observed in adults, though the sample size of adults was limited. Finally, when comparing the three phases of chikungunya described in literature, we observed a strong similarity between interim and chronic arthralgia cases in pediatric cases (≤15 y/o), but not in adult cases (>15 y/o). Our findings provide new insights into chikungunya-associated arthralgia in both pediatric and adult cases that could be used to improve clinical guidelines.

There are limited longitudinal data on the age-specific changes in the risk of chronic arthralgia, particularly in Latin American populations (10–12,15). We observed that a substantial ∼70-95% of pediatric cases reported arthralgia, and of the total arthralgia reported among pediatric cases, ∼20-30% occurred beyond the acute phase, demonstrating that many pediatric cases are vulnerable to long-term arthralgia. Further, we observed a distinct transition between the prevalence of acute and interim/chronic arthralgia around age 30-35, and the transition age decreased to around age 20 when the interim and chronic phases were considered together. Notably, age 20 is far below the threshold of 45 years commonly used in clinical guidelines to demarcate risk for post-acute arthralgia (2,6,13). These observations highlight the burden of chronic, chikungunya-associated arthralgia beginning at a younger age than often described in the literature (6,13,14). Importantly, although quality-of-life research in children with chikungunya is lacking, several studies have demonstrated the negative effect of arthralgia on the livelihood of adult cases (32,33). In a study conducted on La Reunion Island, adult cases reported arthralgia, discomfort, and depression up to six years after acute chikungunya (5). If clinical guidelines do not fully account for the hidden burden of chikungunya in children and young adults, these groups will be at heightened risk of being neglected and suffering negative effects on their livelihood, particularly in resource-limited settings (10,11).

We observed that polyarthralgia in the legs was most pronounced in children, implying that examination of the legs can better serve to diagnose post-acute pediatric arthralgia than other body parts. Further, we note that arthralgia in the elbow clusters with arthralgia in the neck, back, shoulders, and hips, despite the proximity of elbows to the hands and wrists. Little is known about how arthralgia in the joints clusters and manifests post-acutely in children (6,11). Understanding the unique presentation of polyarthralgia among different age groups can facilitate identification of chikungunya, particularly among young children with a limited capacity to detail their pain. The relatively high prevalence of post-acute polyarthralgia suggests long-term follow-up should be conducted for both children and adults.

Our results extend the observed trend that older females (50+ y/o) experience more arthralgia than older males (14,25) to the pediatric population, as we found that pediatric females (0-15 y/o) had significantly higher odds and post-acute hazard of arthralgia when compared to males. Hormonal and immunological differences between females and males has been hypothesized (34) to explain the higher prevalence of arthritic diseases among females, though similarities between chikungunya-associated arthralgia and rheumatoid arthritis are debated (7,35). Altogether, our results and the literature suggest that across all ages, sex is an important risk factor for arthralgia. Consequently, it is critical for clinical guidelines to emphasize the risk and management of chikungunya-associated arthralgia for females of any age.

The WHO recommends that the chronic phase of chikungunya be defined as 12 weeks post-symptom onset (9), with little acknowledgement of the interim phase. Here, we observed distinct trends and presentation of arthralgia in pediatric participants between the first 10 days post-fever onset (acute phase) and thereafter (interim and chronic phases). To date, no analysis has described the prevalence of arthralgia in each phase of chikungunya from childhood to adulthood. Furthermore, the divergence between the acute and interim/chronic phases we observed was particularly evident between age groups. This finding suggests that although interim phase arthralgia in adults is self-limiting, interim phase arthralgia in children differs from adults and can be utilized as an indicator for chronic arthralgia. Vairo et al. (12) and the French Infectious Diseases Society (2) have previously highlighted the similarities between the interim and chronic phases. It has been noted that earlier and more successful clearance of CHIKV during the acute phase results in protection from chronic chikungunya (36,37). Further, early management of acute inflammation has been shown to decrease the risk of chronic inflammation (38). Interim arthralgia might be linked to poor viral clearance and constitutes a higher risk for long-term symptoms. If guidelines explicitly associate pediatric interim arthralgia with greater odds of chronic arthralgia, medical professionals could identify cases with earlier signs of arthralgia as being at risk for prolonged arthralgia and ensure long-term follow-up to protect pediatric cases.

Our study has several limitations. Our results are based in part on reports of polyarthralgia from pediatric patients, which might introduce misclassification bias due to subjectivity when clinically probing for signs of arthralgia. However, our data is supported by physicians that have over 10 years of experience in diagnosing arthralgia and related symptoms in young children due to the high burden of arboviral disease in Managua (26). Further, our study is limited by the small sample size of adult participants, which was by design as we aimed to primarily characterize children to fill gaps in the literature. Finally, the generalizability of our findings may be limited to the Asian CHIKV genotype, due to previously described variability in clinical presentations of chikungunya across lineages (14,22).

Overall, our results provide new insights into chikungunya-associated arthralgia and demonstrate the burden of arthralgia in pediatric cases, both in the acute and post-acute phases. We observe a strong age-prevalence trend for arthralgia and suggest improvements for the pediatric definition of the phases of chikungunya. Overall, our results inform chikungunya clinical guidelines for short-term and long-term care in both pediatric and adult populations.

## Data Availability

All data produced in the present study are available upon reasonable request to the authors

## VI. Acknowledgments

We thank the study participants for taking time out of their lives to help us understand more about the presentation of chikungunya. This study was made possible by the dedicated clinical, laboratory, and data scientists at the Sustainable Sciences Institute and Centro de Salud Sócrates Flores Vivas in Managua, Nicaragua. Their passion for improving our understanding of neglected tropical diseases benefits at-risk communities across the globe. We thank Victoria Warnes for her support. This work was made possible by the R language and RStudio teams that provide flexible and comprehensive open-source data analysis technology. This study was funded by NIH grants P01AI106695 (to E.H.), U19AI118610 (to E.H.), and R01AI099631 (to A.B.).

## Author Contributions

Conceptualization: C.M.W., F.B.C., E.H.; Funding acquisition: E.H., A.B.; Investigation: C.M.W., F.B.C.; Methodology: C.M.W., F.B.C.; Project administration: S.O., A.B., G.K., E.H.; Supervision: F.B.C., E.H.; Visualization: C.M.W.; Data Sourcing: J.V.Z., B.L.; Writing – original draft: C.M.W., F.B.C., E.H; Writing – review & editing: C.M.W., F.B.C., E.H

## Declarations of Interest

The authors declare no competing interests.

## Notes

### Competing Interest Statement

The authors have declared no competing interest.

### Funding Statement

This study was funded by NIH grants P01AI106695 (to Eva Harris), U19AI118610 (to Eva Harris), and R01AI099631 (to Angel Balmaseda).

### Author Declarations

This study was approved by the Institutional Review Boards of the University of California, Berkeley, and the Nicaraguan Ministry of Health.

### Summary of Updates

Figures 4A and 4B were switched. Just a correction on the order.

