## Supplemental Materials for "Longitudinal Analysis of the Burden of Post-Acute Chikungunya-Associated Arthralgia in Children and Adults: A Prospective Cohort Study in Managua, Nicaragua (2014-2019)"

### I. Methods

#### Inclusion criteria

Participants were assessed based on laboratory-confirmed or suspected clinical presentation with chikungunya. Diagnostic criteria used during the 2014-2015 chikungunya epidemics were based on clinical standards for evaluating suspected arboviral infections that circulate in Managua, Nicaragua. These criteria encompass the World Health Organization's case definition for suspected chikungunya cases (1). Suspected chikungunya cases were assessed for 1) undifferentiated fever, or 2) fever or feverishness and >2 of the following: headache, muscle pain, joint pain, retro-orbital pain, rash, hemorrhagic manifestations, and leukopenia (2). Suspected cases were confirmed as CHIKV infections if 1) CHIKV real-time RT-PCR results were positive, 2) seroconversion was observed using an anti-CHIKV immunoglobulin M enzyme-linked immunosorbent assay (ELISA), or 3) seroconversion or a 4-fold or greater increase in anti-CHIKV antibody titers was observed with an in-house inhibition ELISA using paired acute and convalescent samples (3).

Individuals aged 6 months and older were considered for the study. Inclusion criteria for the study were as follows: 1) laboratory-confirmed or clinically probable CHIKV infection, 2) having been attended to at the HCSFV, 3) residing in the HCSFV catchment area during the study period, 4) having provided written informed consent themselves or through a legal representative if <18 years old, and 5) having provided verbal assent if between 6 and 17 years old (3,4). All participants were identified as suspected chikungunya by study physicians, and serum samples were collected to determine the infectious status based on the diagnostic methods described above.

All pediatric patients between 2-14 years old (y/o) were active participants in the Pediatric Dengue Cohort Study and were recruited from the HCSFV upon fulfillment of inclusion criteria (5). Children between 6 months to 2 y/o and adult participants were recruited from HCSFV upon fulfillment of inclusion criteria.

#### Data processing

Participants were included in the analysis even if data were not available for every follow-up visit. Follow-up visit data were considered up to a maximum of 625 days post-fever onset. Participants above the age of 55 were omitted from this study due to low sample sizes beyond age 55. Of the 811 participants in the study, 44 were removed due to disqualifying, missing, or incomplete data, bringing the total participants to 767 individuals. Participants were stratified into age ranges 0-4, 5-9, 10-15, and 16+ based on established protocols within the PDCS and the Nicaraguan Ministry of Health. Adults were grouped into one range, 16+, due to the small sample size that prohibited further stratification. When directly comparing pediatric and adult cases, the pediatric group was condensed to ages ≤15 and the adult group was considered as ages 16+. Arthralgia in the hands, wrists, elbows, shoulders, neck, back, hips, knees, ankles, and wrists was defined as the report of either arthralgia, edema, tenosynovitis, joint abnormality, or difficulty moving, for the purposes of this analysis.

**Zika pediatric cohort**

Data was utilized from a longitudinal pediatric cohort of confirmed and symptomatic Zika virus infections (n=161) in Managua, Nicaragua. Participants were monitored for over ~18 months for signs of arthralgia. The Zika cohort study ran parallel to the chikungunya cohort study and involved the same inclusion criteria and study design. Participants in the Zika cohort study were diagnosed based on established PDCS clinical criteria for suspected Zika and confirmed through PCR methods (6).

**Phases of chikungunya**

Participant visits were classified as acute, interim, and chronic arthralgia cases of chikungunya. The acute cases were defined as experiencing their entire period of illness within the first 10 days post-fever onset based on CDC and World Health Organization (WHO) recommendations (7,8). Acute chikungunya was initially stratified by both ≤15 days and ≤10 days post-fever onset in our analysis, with no measurable difference in results. The chronic cases of chikungunya were defined as experiencing chikungunya-consistent signs or symptoms ≥90 days post-fever onset based on WHO and French guidelines (8,9). The interim cases experienced illness during the period from 11-89 days post-fever onset (9,10). Collectively, the interim and chronic cases constitute the post-acute phase of chikungunya, which we define as any reported arthralgia 10 days post-fever onset. For our analysis, participants who experienced arthralgia in multiple chikungunya phases were analyzed by the last phase in which they reported arthralgia, leading to the designation of individuals as an acute, post-acute, interim, or chronic "arthralgia case". This terminology was used to distinguish individuals by the length of their chikungunya-associate arthralgia symptoms.

**Statistical analysis**

We used logistic regression models to estimate odds ratios (OR) for binary outcomes, such as the presence or absence of arthralgia. We considered both unadjusted models and models adjusted for sex and age. In our analysis modeling the relationship between the three phases of disease (*i.e.,* acute, interim, chronic phases), we adjusted for age and sex as covariates in the model, based on supporting evidence from the literature and our own analyses (10). General additive models (GAM) – semi-parametric extensions of traditional models – were used to visualize the non-linear relationship between different variables and were preferred over generalized linear models (GLM) due to the addition of smoothing functions that better estimate the functional relationships between outcome and explanatory variables. GAMs relax the assumption of linearity, which allows for the non-linear estimation of trends (11). Pearson's Chi-squared test was used to compare the proportion of participants with arthralgia stratified by different criteria (12,13).

Survival analyses were used to describe the percent of participants reporting arthralgia over time, accounting for censoring. Survival data were visualized using Kaplan-Meier (KM) curves, which plot the probability/percent of the event not occurring across the study period (14). KM survival curves were evaluated using the log-rank test, with the null hypothesis proposing that the plotted survival curves are similar. We evaluated group trends in the KM survival curves using the log rank test for trend (15). We omitted confidence intervals in the visualization of the KM survival curves to improve graphical presentation and use the p-value from the log-rank test to represent the statistical differences between the curves. Weibull survival models were used to calculate the hazard ratio of experiencing the outcome over the study period, compared to a reference group (15). Survival analysis was utilized only when observing the data beginning from the interim phase of disease (>10 days post-fever onset) until study cessation. Hazard ratios were not calculated for the acute phase of disease because the proportional hazard assumption was not satisfied during this phase. Survival analyses were conducted using the survival (v3.2-13), survminer (v0.4.9), and eha (v2.9.0) packages and visualized using base R plotting.

Agglomerative hierarchical clustering was used to construct a co-occurrence dendrogram of participants' signs and symptoms (16). The Ward method was used to create groups with minimal variance, and the Manhattan distance was used to determine the underlying distance matrix. The cophenetic distance correlation coefficient was calculated to measure the similarity between the original distance and the cophenetic distance (16). The higher the cophenetic distance correlation coefficient is, the more appropriately the dendrogram represents a hierarchical structure present in the original data. Analyses were conducted using the base stats package, and the dendrograms were visualized using base R graphics and the dendextend (v1.15.2) package.

For sensitivity analysis, we used KM estimates to evaluate any differences in reported arthralgia occurrence between the PCR-confirmed and clinically/epidemiologically probable groups, Further, to evaluate whether the baseline arthralgia among pediatric individuals might confound our results, we conducted a comparison between our results and a Zika pediatric cohort. We compared the reported arthralgia occurrence beyond the acute phase of disease (>10 days post-fever onset) using KM estimates. Our comparison of the two cohorts was conducted with the consideration that Zika manifests similarly to chikungunya in the acute phase of disease, but it has not been shown to contribute to chronic arthralgia (17,18).

Data were analyzed using R language version 4.1.1 within the RStudio (2021.09.0, Build 351) integrated development environment. Data manipulation was conducted using the core tidyverse (v1.3.1) packages along with base R (v4.1.1), lubridate (v1.8.0), reshape2 (v1.4.4), broom (v0.7.9), epitools (v0.5-10.1), plyr (v1.8.6), and tableOne (v0.13.0) packages. Two-dimensional plots were visualized using the base R graphics and ggplot2 (v3.3.5) packages, supplemented with the scales (v1.1.1) package to express percentages. Tables were created using Microsoft Word 2021 (v16.56).

### II. Supplemental Tables

**Supplemental Table 1. Prevalence of reports of arthralgia in the acute and post-acute phase**

| Age Range | Acute (%) | Post-Acute (%) | Total |
|---|---|---|---|
| **0-4** | 73 (73.7) | 26 (26.3) | 99 |
| **5-9** | 158 (67.8) | 75 (32.2) | 233 |
| **10-15** | 275 (68.8) | 125 (31.3) | 400 |
| **16+** | 124 (56.6) | 95 (43.4) | 219 |

[a] Some participants were omitted due to missing acute stage data, n = 41

**Supplemental Table 2. Proportion of visits with reports of arthralgia**

| Age Range | Number of visits with reported arthralgia[a] | | | | | |
|---|---|---|---|---|---|---|
| | 0 | (%) | 1 | (%) | 2+ | (%) |
| 0-4 | 23 | (22.5) | 60 | (58.8) | 19 | (18.6) |
| 5-9 | 30 | (15.2) | 122 | (61.6) | 46 | (23.2) |
| 10-15 | 22 | (8.3) | 182 | (60.3) | 98 | (32.5) |
| 16+ | 2 | (1.6) | 55 | (43.7) | 70 | (55.1) |

[a] Some participants were omitted due to missing acute stage data, n = 41

**Supplemental Table 3. Hazards for chikungunya-associated arthralgia**

| Age range | Hazard ratio (95% CI) | |
|---|---|---|
| 0-4 | 0.51 | (0.25, 1.07) |
| 5-9 | 0.74 | (0.43, 1.27) |
| 10-15 | - | |
| 16+ | 3.72 | (2.28, 6.05) |
| **Sex** | | |
| Female | 2.27 | (1.49, 3.46) |
| Male | - | |

[a] >10 days post-fever onset

**Supplemental Table 4. Prevalence of post-acute polyarthralgia**

| Category | Reported Arthralgia | | | |
|---|---|---|---|---|
| | **No** | **(%)** | **Yes** | **( %)** |
| All ages [b] | | | | |
| Legs | 549 | (71.5) | 219 | (28.5) |
| Hands | 605 | (78.8) | 163 | (21.2) |
| Torso | 664 | (86.5) | 104 | (13.5) |
| Pediatric ($\leq$ 15) [b] | | | | |
| Legs | 467 | (76.3) | 145 | (23.7) |
| Hands | 515 | (84.1) | 97 | (15.9) |
| Torso | 551 | (90.0) | 61 | (10.0) |
| Adult (16+) [b] | | | | |
| Legs | 84 | (52.9) | 73 | (47.1) |
| Hands | 91 | (58.0) | 66 | (42.0) |
| Torso | 114 | (72.6) | 43 | (27.4) |

[b] All categories are significantly different, p-value < 0.00

**Supplemental Table 5. Prevalence of each arthralgia case by age**

| | Acute (%) | | Interim (%) | | Chronic (%) | | No Pain (%) | |
|---|---|---|---|---|---|---|---|---|
| 0-4 | 55 | (53.9) | 7 | (6.9) | 17 | (16.7) | 23 | (22.5) |
| 5-9 | 113 | (57.1) | 16 | (8.1) | 39 | (19.7) | 30 | (15.2) |
| 10-15 | 179 | (59.3) | 30 | (9.9) | 71 | (23.5) | 22 | (7.3) |
| 16+ | 55 | (43.3) | 33 | (26.0) | 37 | (29.1) | 2 | (1.6) |

[a] Some participants were omitted due to missing acute stage data, n = 41

**III. Supplemental Figures**

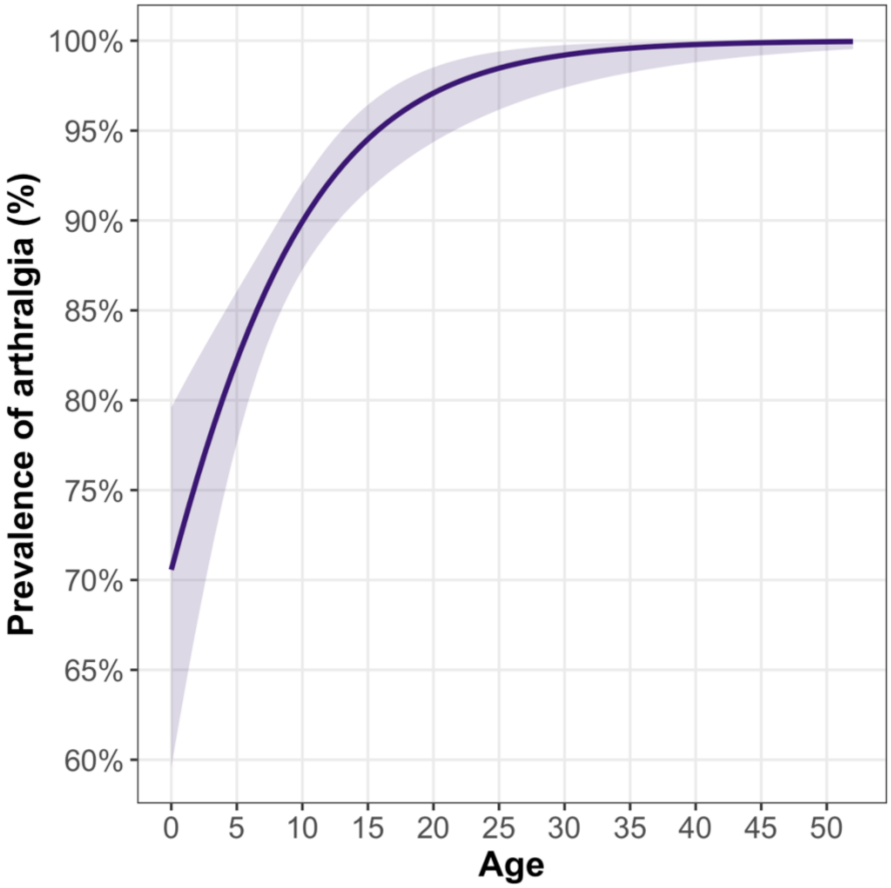


**Supplemental Figure 1. Age trends of prevalence of arthralgia depicted using a GAM model.** A 95% confidence band is shown around the model trend.

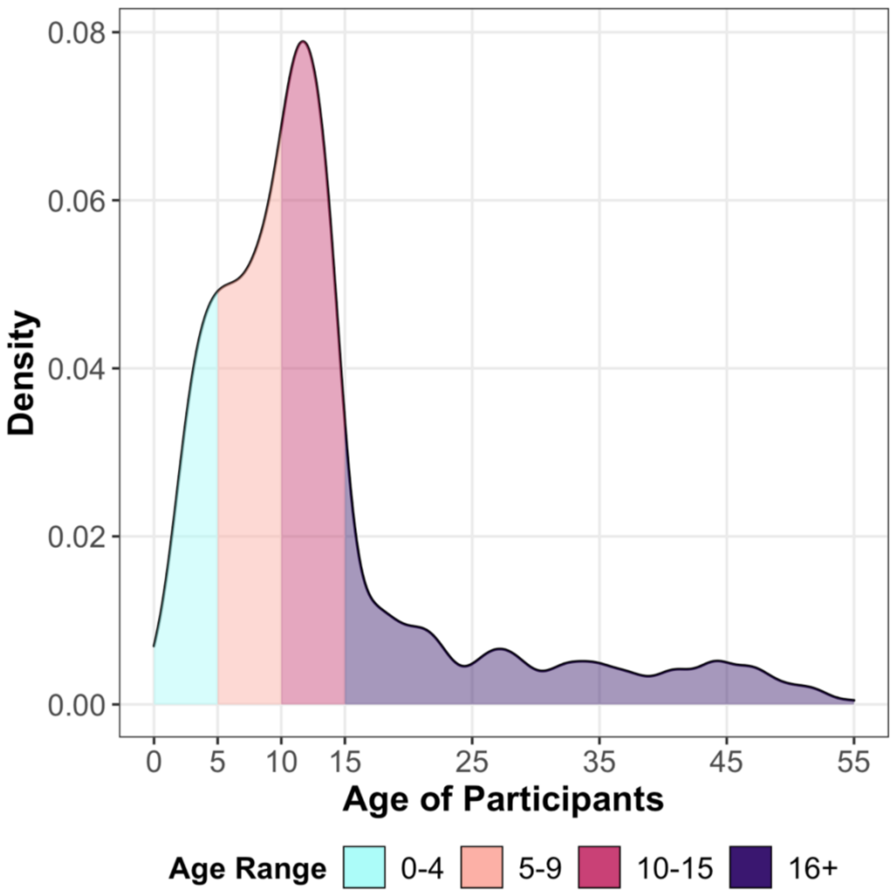


**Supplemental Figure 2. Age density plot of participants.** The percent of participants in this study across age are depicted using an age-density plot. Colors correspond to the age-ranges defined within the study (0-4, 5-9, 10-15, and 16+ years old)

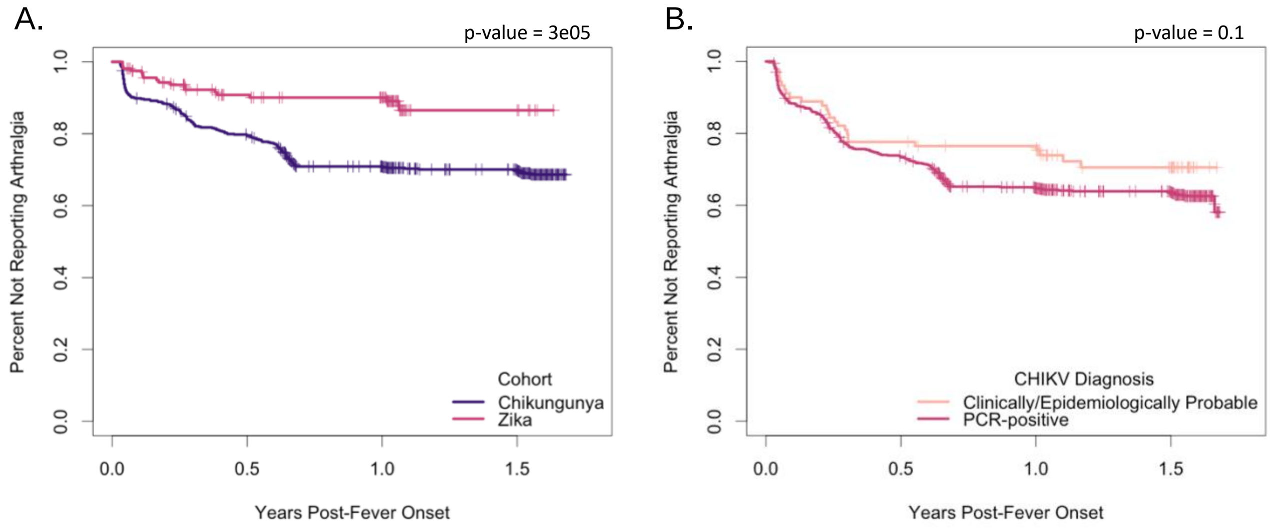


**Supplemental Figure 3. Kaplan-Meier plot demonstrating percent of participants not experiencing arthralgia by diagnostic method and cohort.** Ticks correspond to censoring events. Panels show the distribution of participants beginning 10 days post-fever onset and ending at the last reported data point stratified by diagnostic method **(A)** or cohort **(B)**. We observed no differences in the presentation of arthralgia beyond the acute phase of disease between the RT-PCR-positive and clinically/epidemiologically probable groups (A). We observed a significant difference in the presentation of arthralgia beyond the acute phase of disease between the pediatric participants in the Zika and chikungunya cohorts (0-15 y/o) (B).
